# Prognostic significance of AI-identified markers for isoniazid resistance in Mycobacterium tuberculosis

**DOI:** 10.1101/2023.11.29.23299121

**Authors:** Siavash Valafar, Aram Valafar

## Abstract

Antibiotic resistance in tuberculosis (TB), a disease that kills 1.5 million people annually, is a great concern. The emergence of drug resistance in *M. tuberculosis*, the obligate pathogen of TB seems to follow an order. In most cases resistance to isoniazid (INH) emerges first, followed by rifampicin, then either pyrazinamide or ethambutol, and finally followed by resistance to second-line drugs. Prevention of emergence of INH resistance can go a long way to prevent the emergence of resistance to other drugs. In this manuscript we present the prognostic value of specific mutations in the hope that resistance can be e predicted and hence avoided. Here we present evidence that resistance to INH follows a stepwise evolutionary trajectory in most cases. This information can therefore be used to predict and avoid INH resistance. In our approach, we used genomic and phenotypic data from over 16,000 samples collected by two large databases, the TB Portals and the CRyPTIC consortium. We used a deep learning neural network model to identify promising mutations using the TB Portal data. We then tested the prognostic value of the identified mutations using the CRyPTIC consortium data. In this manuscript, we estimate a prognostic accuracy of 73% for correctly predicting the emergence of three canonical INH resistance mutations (katG315, inhA-15, and inhA-8) by using two prognostic markers. Additional time course samples and analysis will undoubtedly uncover prognostic markers for other evolutionary trajectories that lead to resistance.

## 1. Introduction

*Mycobacterium tuberculosis* (*M. tuberculosis*), the causative agent of the tuberculosis (TB) disease, infects millions annually.^1^ Annually, 10.5 million people develop the active form of the disease for the first time, globally.^1^ TB is commonly treated by the standard regimen recommended by the World Health Organization (WHO) which includes the combination of the four “first-line” drugs ethambutol (EMB), isoniazid (INH), rifampicin (RIF), and pyrazinamide (PZA).^2^ Unfortunately, the emergence of antibiotic resistance is a major challenge in global TB control.^1^ The WHO estimates 1.3 million people developed isoniazid resistant (INH^R^) TB, globally.^1^ This included an estimated 410,000 cases of multidrug resistant tuberculosis (MDR-TB) where the bacterial strains are resistant to both RIF and INH.^1^ Treatment of MDR-TB is challenging and estimated to succeed only in 52% of cases (without new and repurposed drugs [NRDs]).^3^ Yet more complex is the treatment of extensively drug resistant tuberculosis (XDR-TB) cases where the strains are MDR-TB but also resistant to any fluoroquinolone and at least one additional Group A drug. Treatment of XDR-TB cases is only successful in less than 25% of cases without NRDs (bedaquiline, delamanid, pretomanid, clofazimine, and linezolid). While NRDs have offered renewed hope and significantly improved the success rate of MDR-TB and XDR-TB treatment (between 60%^4^ and 89%^5^ for MDR and 48.6%^6^-90%^7^ for XDR-TB in some trials with the six-month BPaL or BPaLM regimens), the cost of these drugs, their availability, and increasing incidence of resistance to them in programmatic application globally is a distinct concern.

Emergence of antibiotic resistance generally follow an order where INH resistance emerges first, followed by RIF resistance, followed by resistance to PZA or EMB, and finally to second line drugs.^8–13^ It has been hypothesized that prevention of emergence of INH resistance increases the likelihood of sterilization and reduces the likelihood of progression to MDR-TB^14–17^ through timely increased INH dosing or replacement with another drug alternative. Three mutations in the *M. tuberculosis* genome are responsible for an estimated 81.4% of all reported INH^R^ cases globally.^18^ By far the most common INH resistance-causing mutation is in codon 315 of the catalase peroxidase gene *katG* (*katG*315), followed by the -15 and -8 mutations in the promoter of the operon that includes the *inhA* gene (*inhA*-15, *inhA*-8).^18^ In this article we explore the potential of using genomic markers in *M. tuberculosis* for prognostic purposes. Specifically, we ask whether any genomic markers in *M. tuberculosis* can predict the emergence of these three INH resistance-casing mutations. We used a deep learning artificial intelligence (DL-AI) model to identify two markers for this prognostic purpose. We then used the Cryptic consortium genome collection to estimate the prognostic value of the two mutations.

## 2. Methods

### Data

In total 11,747 sets of whole genome sequencing (WGS) and associated INH drug susceptibility testing (DST) results were included in this study from two publicly available sources:

a. *TB Portal*: The first set included 4,131 clinical isolates from TB Portals^19^ for training the DL-AI model. Of these, we selected for inclusion 1,931 genomes and their associated INH DST collected from 453 patients. The selection criteria for these were that each patient had to have at least two time-course samples that 1) one sample was from prior to their treatment start (baseline sample) and at least one was from after treatment start, 2) each had associated INH DST data, and 3) had WGS data available for the nine INH-resistance associated genes: *katG, inhA, ndhA, fabG1, ahpC, Rv1258c, mshA, Rv2752c*, and *ndh*.
b. *CRyPTIC Consortium*: The second dataset included 12,289 clinical isolates from the CRyPTIC consortium^20^ which was used for the assessment of the prognostic sensitivity of the DL-AI identified markers. Of the total of 12,289 samples stored in the CRyPTIC database, we included 9816 genomes from 919 patients. The selection criteria for these were the same as those for the TB Portals samples.

### Patient Categorization

Patient groups from both sets were divided into three categories:

a. EMERGED: group of patients for whom the emergence of at least one of the three canonical mutations (*katG*315, *inhA*-15, *inhA*-8) were observed during their treatment.
b. INITIAL RESISTANCE: group of patients for whom at least one of the canonical mutations was observed in the baseline sample.
c. NOT EMERGED: group of patients for whom none of the three canonical mutations were never observed in any of their samples.

### Deep Learning (DL)

In total the genomic and phenotypic data from 453 patients from TB Portal to train the DL model. The goal of this exercise was to identify the mutations that carried the highest prognostic power for prediction of emergence of the three canonical mutations (*katG*315, *inhA*-15, *inhA*-8). For each patient, two sets of samples were prepared.

#### Input data

Mutations in the listed nine genes from the baseline sample of each patient were provided to the models as binary input data. In total 49 mutations were identified in these genes across all patients (Supplementary Table ST1). For each patient a binary vector of presence/absence vector (length 49) was generated based on the patient’s baseline genomic data.

#### Desired target data

The last sample of the patient was evaluated for the presence of the three canonical mutations. For patients whose follow up samples (any sample other the baseline sample) contained any of the canonical mutations, a desired target value of 1 was designated, indicating the emergence of the canonical mutations. If none of the canonical mutations were observed for any of the time-course samples of the patient, a value of 0 was designated as the desired target.

#### Model topology, training, and experimental design

In this study, we only explored the prognostic value of up to two mutations. As such, a Deep Neural (DN) model was trained for each of the 49 mutations individually, and one was trained for any combination of two mutations from the set of 49 (1,176 combinations). As a result, the total number of trained models was *49C1* + *49C2* = 1,225.

All models had a three-layer topology with two hidden layers and one output layer. Size of the three layers for the single-mutation models were optimized empirically to be 3-2-1. While layer optimization for the two-mutation models resulted in 5-3-1 networks. For each model a 10-fold cross-validation process was used to evaluate the performance of the model. We split the 453 patients into a training set (including cross-validation) of 250 randomly selected patients and a testing set of 203 patients. The random selection was limited to patients within each of the three categories of EMERGED, INITIAL RESISTANCE, and NOT EMERGED sets to ensure that each of these categories were properly represented in both the training and the testing sets. The trained models were then used to predict the emergence of any of the three canonical mutations in the testing set. For these experiments the Deep Learning/Statistics and Machine Learning toolboxes of MATLAB version R2023a were used.

### Sensitivity assessment

The CRyPTIC data was used to assess the prognostic sensitivity. The mutations that produced the best performing DN models were selected for the sensitivity assessment. This was done for all individual mutations and the top performing combination of two mutations. For each individual mutation and the best performing combination of two mutations, we assessed the sensitivity as the ratio of the correctly prognosed patients to the total number of patients in the CRyPTIC data set.

## 3. Results

### Machine Learning results

The mutations identified as most effective in predicting the emergence of the three canonical mutations as well as the prediction accuracy of the model that used each of these mutations, individually, are shown in Table 1. In this exercise 49 sets of models were built, one for each mutation. The accuracies reported in Table 1 are the highest achieved accuracy in each set. As it can be seen, the best performing model used the mutation *Rv1258c* 581 indel and was able to reach a predictive accuracy of 66.33%.

**Table 1:** List of mutations that were best predictive of the emergence of the canonical mutations (*katG315, inhA-15, inhA-8*). Note: Two mutations, mshA N111S and katG R463L.

| Mutation | Accuracy |
| --- | --- |
| ***Rv1258c* 581 indel** | 66.33% |
| *mshA* A187V | 60.00% |
| *katG* A106V | 53.00% |
| *ndhA* M325T | 52.33% |
| *ndhA* 371V | 51.00% |
| *Rv2752c* A520A | 51.00% |
| *fabG1* S144S | 47.00% |
| *ndhS* 374S | 45.00% |

Of note is that in this exercise we excluded the mutation *katG* R463L since it is a known genetic background mutation and is roughly equally prevalent among resistant and sensitive isolates. Also, because DL models are capable of exploiting inverse relationships for their predictions, they can exploit mutations that predominantly appear in cases that remain sensitive throughout the therapy for their predictions. However, because the focus of this study was to identify the markers that emerge in eventually resistant cases, we did not include mutations that appeared in a few resistant cases but in many sensitive cases. An example of this is the mutation *mshA* N111S which predominantly appeared in sensitive isolates. This mutation provided increased accuracy but was excluded due to the goals of this project, as mentioned. (Please see discussions).

To further improve the prediction accuracy, we built models for every combination of two mutations from the list of 49 (Supplementary Table 1). This produced 1,176 (*49C2*) sets of models, each using two distinct mutations from the list of 49 (Supplementary Table ST1). The highest accuracy of all 1,225 DL models is shown in Figure 1. The highest prediction accuracy (77.33%) belonged to the model trained with the combination of two mutations: *ndhA* M325T and *Rv1258c* 581 indel. The second-best performing combination was the combination of *Rv1258c* 581 indel and *mshA* A187V mutations with an accuracy of 71.33%.

**Figure 1:**
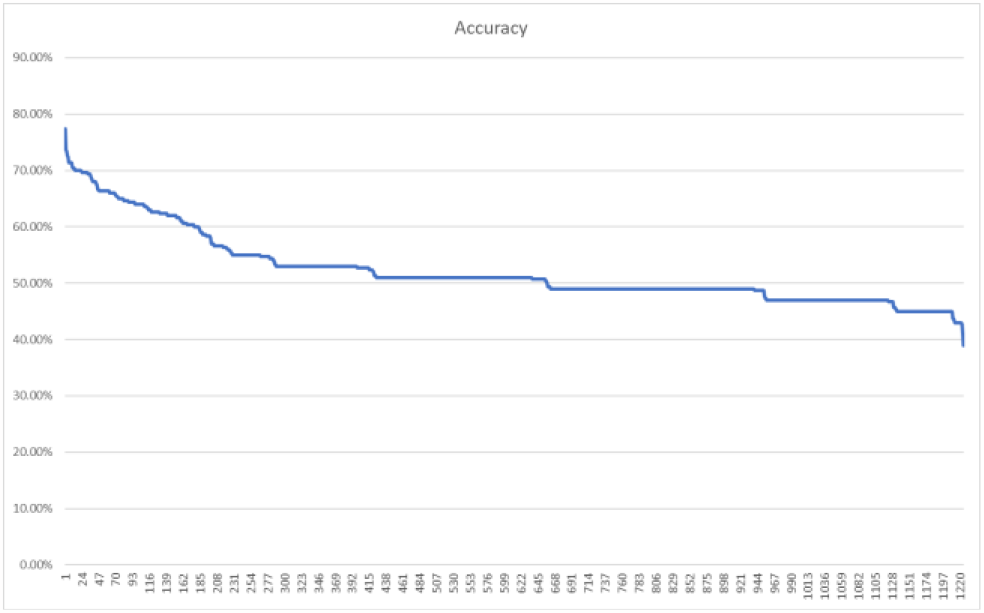
Deep Neural model accuracy in predicting the three canonical mutations (*katG*315, *inhA*-15, *inhA*-8) using at most two of the list of 49 candidate mutations (i.e. _*49*_*C*_*1*_+_*49*_*C*_*2*_ = 1,225 combinations).

### Validation of the DL results

In order to validate the results of the DL models, we used 9816 additional genomes from 919 patients from the CRyPTIC consortium^20^ that was not included in training the DL models. The selection criteria for these were that each patient had to have at least two time-course samples that 1) each had associated INH DST data, and 2) had WGS data available for the nine listed genes. Of the 919 patients, 27 did not have any of the three canonical mutations in their baseline sample but experienced the emergence of at least one of the three mutations during their treatment. We refer to this group as the EMERGED Group. An additional 330 patients had at least one of the canonical mutations in their baseline sample. We refer to this group as the INITIAL RESISTANCE group. Finally, in 562 patients the three canonical mutations never emerged during treatment. We refer to this group as the NOT EMERGED group. These were mostly phenotypically INH susceptible patients. Table 2 displays the breakdown of the 27 patients in the EMERGED category based on the observation of the two precursor mutations *Rv1258c* 581 indel and *mshA* A187V.

**Table 2:** The breakdown of the 27 patients in the EMERGED group based on observation of the two precursor mutations *Rv1258c* 581 indel and mshA A187V prior to the three canonical resistance mutations (*katG315, inhA-15, inhA-8*).

| Precursor | Patient Count |
| --- | --- |
| *Prior observation of both* | 7 |
| *Concurrent observation of both* | 6 |
| *Concurrent observation of Rv1258c 581 indel* | 4 |
| *Concurrent observation of mshA A187V* | 1 |
| *Prior observation of Rv1258c 581 indel* | 1 |
| *None observed* | 7 |
| *Superinfection (excluded from this study)* | 1 |

In summary, in 19 of the 27 patients (73%) that experienced emergence of at least one of the three canonical mutations, at least one of the two precursor mutations *Rv1258c* 581 indel and *mshA* A187V was observed. Of the 19 patients, in seven, the two precursor mutations were observed in a time point prior to the observation of the canonical mutation. In another six the two precursor mutations were observed at the same time point when the canonical mutation was observed. The great majority of these patients only had samples from two timepoint. In one more patient *Rv1258c* 581 indel was observed prior to the emergence of a canonical mutation, and in one other *mshA* A187V was observed concurrently with the canonical mutation for the first time. This patient only had two samples available. In 7 patients (17%) a canonical mutation emerged without a prior or concurrent occurrence of the precursor mutations. Finally, one patient seemed to have undergone superinfection with the isolate from the second timepoint being different from the baseline. This patient’s data was excluded from all further analysis.

## 4. Discussions

While new drugs infrequently are developed to combat tuberculosis, the emergence of resistance significantly reduces the shelf life of such drugs, further reducing the appetite of the pharmaceutical companies for antibiotic drug development. Prevention of emergence of resistance, therefore, must remain a priority for the medical research community. Since INH resistance often emerges first in a dark sequence of continued acquisition of additional resistance, prognosis of INH resistance carries a special significance in the battle against drug resistant tuberculosis.

In this manuscript we presented an approach to identify and assess the prognostic significance of promising mutations. Specifically, in this manuscript, we focused on the prognostic significance of the two most promising precursor mutations *Rv1258c* 581 indel and *mshA* A187V. These two mutations were observed in 73% of the isolates collected from patients who experienced emergence of at least one of the three canonical mutations during treatment. In 89% of the patients who had more than two-time course samples and experienced the emergence of the canonical mutations, at least one of the two precursor mutations was observed. The number of such patients was small, even in the large CRyPTIC data set. The great majority of patients only had one sample followed by the group of patients with two samples. The number of patients who had three or more-time course samples was exceedingly small.

In this project, we only considered the prognostic significance of up to two mutations at a time. This was solely done in consideration of the exponentially increasing computational complexity with increasing combinatorial complexity (NP-complete class of problems). As such, this project only exposes the most prominent prognostic markers for the most prominent evolutionary trajectory to emergence of the three canonical mutations. Given these significant limitations, the prognostic sensitivity of 73% is quite promising. Further experimentation that removes some of these limitations, such as consideration of at least three mutations at a time is expected to improve the sensitivity to levels acceptable for clinical implementation.

## Data Availability

All data is available within the manuscript.

